# “#LongCOVID affects children too”: A Twitter analysis of healthcare workers’ sentiment and discourse about Long COVID in children and young people in the UK

**DOI:** 10.1101/2022.07.20.22277865

**Authors:** Sam Martin, Macarena Chepo, Noémie Déom, Ahmad Firas Khalid, Cecilia Vindrola-Padros

## Abstract

**Aims:** With a social media analysis of the discourse surrounding the prevalence of Long COVID in children and young people (CYP), this study aims to explore healthcare workers’ perceptions concerning Long COVID in CYP in the UK between January 2021 and January 2022. This will allow to contribute to the emerging knowledge on Long COVID and identify critical areas and future directions for researchers and policymakers.

**Design:** A mixed methods approach with a discourse, keywords, sentiment, and image analysis, using Pulsar™ and Infranodus.

**Setting:** A discussion of the experience of Long COVID in CYP in the UK shared on Twitter between 1 January 2021 and 31 January 2022.

**Participants:** A sample of health workers with Twitter accounts whose bio has them identifying themselves as HCWs.

**Results:** We obtained 2588 tweets. HCW were responsive to announcements issued by authorities regarding the management of COVID-19 in the UK. The most frequent feelings were negative. The main themes were uncertainty about the future, policies and regulations, managing and addressing COVID-19 and Long COVID in CYP, vaccination, using Twitter to share scientific literature and management strategies, and clinical and personal experiences.

**Conclusions:** The perceptions described on Twitter by HCW concerning the presence of Long COVID in CYP appear to be a relevant and timely issue and responsive to the declarations and guidelines issued by health authorities over time. We recommend further support and training strategies for health workers and school staff regarding the manifestations and treatment of Long COVID in CYP.

**Strengths and limitations of this study:**

– Our online analysis of Long COVID contributes towards an emerging understanding of reported experiential, emotional and practical dimensions of Long COVID in CYP specifically, as well as questions of vaccine hesitancy in CYP with Long COVID.
– We identify key policy areas that need considered attention and focus, such as: a) the provision of psychosocial support with access to quality mental health resources to alleviate the impact that Long COVID can have on the mental health of CYP; and b) the development of clear Long COVID pandemic recovery policies that are informed from a health equity perspective and how this affects CYP living with Long COVID.
– This is one of few studies to collect healthcare workers’ perceptions regarding Long COVID in CYP in the UK, using information from Twitter.
– This study is limited to the perception of those who identified as healthcare workers via their online biographies, and so is not representative of the general UK or the global population.

## INTRODUCTION

More than two years after the WHO declared the outbreak of the new coronavirus a Public Health Emergency of International Concern,^1^ it has been estimated that COVID-19 cases have exceeded 533 million worldwide, claiming the lives of nearly 6.3 million people.^2^ One phenomenon that has generated concern in the population^3^ is that people who have had COVID-19 experience long-term effects (weeks, months or years) from their infection, a phenomenon known as post-COVID conditions (PCC) or LongCOVID.^4^

There is increasing information regarding the clinical manifestation of this condition, particularly in the adult population. The prevalence has been estimated at approximately 43%, being more frequent in individuals hospitalised during acute COVID-19 infection.^5^ While Children and Young People (CYP) have a low likelihood of severe COVID-19 infection,^6^ the information available to date indicates that the presence of Long COVID in this group may be as disabling as in adults, reaching a prevalence ranging from 1% to 51%.^7^

A recently agreed definition indicates that Long COVID in CYP is a condition that occurs when one or more physical symptoms persist for a minimum of 12 weeks after initial positive testing and whose symptoms cannot be attributed to an alternative diagnosis.^7^ Long COVID strongly impacts daily functioning and can develop or continue after COVID-19 infection and may fluctuate or relapse over time.^4,7^

Among the symptoms most frequently attributable to Long COVID in CYP have been fatigue and headache.^7^ In CYP, sleep disturbances, difficulty concentrating, abdominal pain, myalgia or arthralgia, persistent chest pain, stomach pain, diarrhoea, heart palpitations and skin lesions have been added to the symptoms.^8^ One of England’s most significant studies is the CLoCk Study.^9^ This national research matched longitudinal and cohort study in adolescents aged 11-17 years and found the presence of at least three symptoms in 30.3% of adolescents who tested positive at baseline, a higher prevalence for those aged 15-17 years versus those aged 11-14 years.^9^ As research on the treatment and prevalence of Long COVID in CYP at this stage of the pandemic continues to emerge, it is essential to add to the literature by developing studies that determine the condition’s impact on this group.

A powerful tool used during the pandemic has been Twitter. With more than 229 million daily active users, Twitter has become one of the most important social platforms in the world. ^10^ People used Twitter during the COVID-19 pandemic for different purposes: world leaders communicated with citizens, ^11,12^ organisations monitored movement, ^13^ scientists studied public discourse around the pandemic, 14,15 performed sentiment analysis, ^16–18^ and more. In the case of physicians and healthcare workers (HCW), Twitter has been used to share and evaluate scientific evidence, guidelines, technical advice, ^19–21^ and track the course and burden of disease, ^22^ among others.

Using the social media monitoring platform Pulsar,^23^ we aimed to explore healthcare workers’ perceptions concerning Long COVID in CYP in the UK between January 2021 and January 2022. We aim to contribute to the emerging knowledge on Long COVID in CYP and identify critical areas and future directions for researchers and policymakers.

## METHODS

Data for this study was analysed by conducting a Collaborative and Digital Analysis of Big Qualitative Data in Time Sensitive Contexts (LISTEN).^24^ This mixed methods analysis consisted of iterative cycles intercalating team discussion and the use of digital text and discourse analytics tools to analyse related social media data.^24^ We used the LISTEN method to perform quantitative and qualitative analyses of Twitter posts, extracted through the Pulsar Platform™, ^23^ related to the experience of Long COVID in CYP in the UK (phrases, words, hashtags, videos, images), published between 1 January 2021 and 31 January 2022. We created an advanced boolean search for keywords mentioning Long COVID and co-related words, hashtags, and symptoms - we also filtered for user accounts who identified as HCW in their Twitter Bio description (Appendix 1).

Quantitative analysis of all tweets included:

i) engagement analysis, where specifically, reactions to posts were measured, e.g., a retweet, a share, or a comment/quote made towards a tweet; ii) sentiment and emotion analysis: an overview of the positive or negative sentiment measured in the words and tone of each post, within the context of Long COVID and HCW’ roles (Appendix 3); iii) emotion analysis, where we measured the emotions expressed in the tweets, classified as sadness, anger, disgust, fear and joy; iv) frequency analysis - looking at the frequency of keywords and themes in the dataset; v) segmentation analysis, a measurement of the key the connections or relationships between keywords and their frequent use in the same context; vi) demographic analysis - where the occupation, gender, city of origin related to the users posting tweets was measured. In addition, and finally, vii) analysis was made of the most influential accounts and the most mentioned websites).

Big qualitative analysis was carried out through thematic discourse analysis of the data sample, using Infranodus,^25^ specifically analyse the key themes and topics of concern expressed throughout the dataset. A codebook was constructed based on the three researchers’ agreed mapping of themes (Appendix 2).

## RESULTS

### Audience analysis

During the period January 2021 to January 2022, we obtained 2588 tweets, coming from 936 accounts. By gender, 32.9% were female, 23.5% male and 43.6% unknown. According to the description given in user’s biography, the most frequent self-reported terms were “NHS” (22.5%), “health” (8.9%), “medical” (6.5%), “nurse” (6.4%), “clinical” (6.2%), “mum” (6.1%), “doctor” (5.6%) and “gp” (5.6%). By city, tweets came mainly from London (37%; n=469), Newcastle upon Tyne (12.6%; n=160), Redcar (6.2%; n=78), Manchester (5.4%, n=69) and Bradford (4.3%, n=54).

### Profession

By profession described in the bio, the most frequently mentioned roles were nurses (6.83%; n=80), medical roles, e.g., paramedic, nursing assistant (6.7%; n=78), clinical, e.g., surgeon, physiotherapist, anaesthesiologist (6.2%; n=73), GP, e.g., hospital GP or local surgery GP (5.5%; n=64), and doctor (5.4%; n=63). The most frequent organisation affiliated with was the NHS (22.7%; n=266).

### Most Influential Accounts

One of the accounts that generated the highest number of mentions, and therefore some of the most influence as they were the ones who talked the most about Long COVID in CYP was the account for @longcovidkids (22.9%; n=211 tweets), related to the most shared website *longcovidkids*.*org*,^26^ an international UK-based charity for families & children living with Long COVID. Although the account was created in October 2020, it was first mentioned in our data collection timeline on 01/01/2021. It offers online support services, funding, and research participation, and representing CYP living with Long COVID in expert forums, research panels, health organisations, and parliamentary groups. The other most shared web pages were theguardian.com^27^ (UK), bbc.co.uk^28^ (UK), peoplewith.com^29^ (US), and ncbi.nlm.nih.gov^30^ (US). This shows that in the UK, there was a mixed influence of UK and US link resources linked to by HCW Twitter users in the UK.

### Keyword analysis

The volume of social media engagement in the discussion about the Long COVID experience in CYP in the UK reached 1,400 posts, 1,550 engagements, and 1.9 million impressions. Overall, comments were very responsive to government decisions regarding the vaccination program and school closures (Appendix 1: Table 1). During the first peak of comments in January 2021, the amount of discourse expanded leading up to March 2021, when there were different announcements of school closures, and the guidelines were delivered regarding the priority groups of the vaccination program (frontline HCW and over 80 years of age first). The highest engagement was between June and July 2021, which coincides with the government announcement regarding the availability of vaccines for people over 18 years of age. The third peak of comments occurred in September 2021, the same month the authorities announced the extension of the vaccination program to children between 12 and 15 years old.

### Top Keywords analysis

The top words associated with CYP’s experience of Long COVID in the UK were: ‘Children’ (13.6%; n=1144), ‘kids’ (6.2%; n=524), ‘people’ (6.0.9%; n=505), ‘Young’ (5.7%; n=485) and ‘schools’ (3.2%; n=273). The top hashtags were #longcovid (53.6.1%; n=738), #longcovidkids (17.3%; n=238), #covid19 (14.3%; n=197) and #covid (6.8%, n=93).

### Sentiment and emotions analysis

According to sentiment (Figure 1), 99.4% of the words included in the word cloud reflected negative sentiments and 0.6% with positive sentiments. Negative sentiments were mainly associated with comments on hospitalisation figures related to COVID-19, criticism of pandemic mitigation policies, and vaccination of CYP. In addition, positive feelings related to the support provided by community support groups.

**Figure 1:**
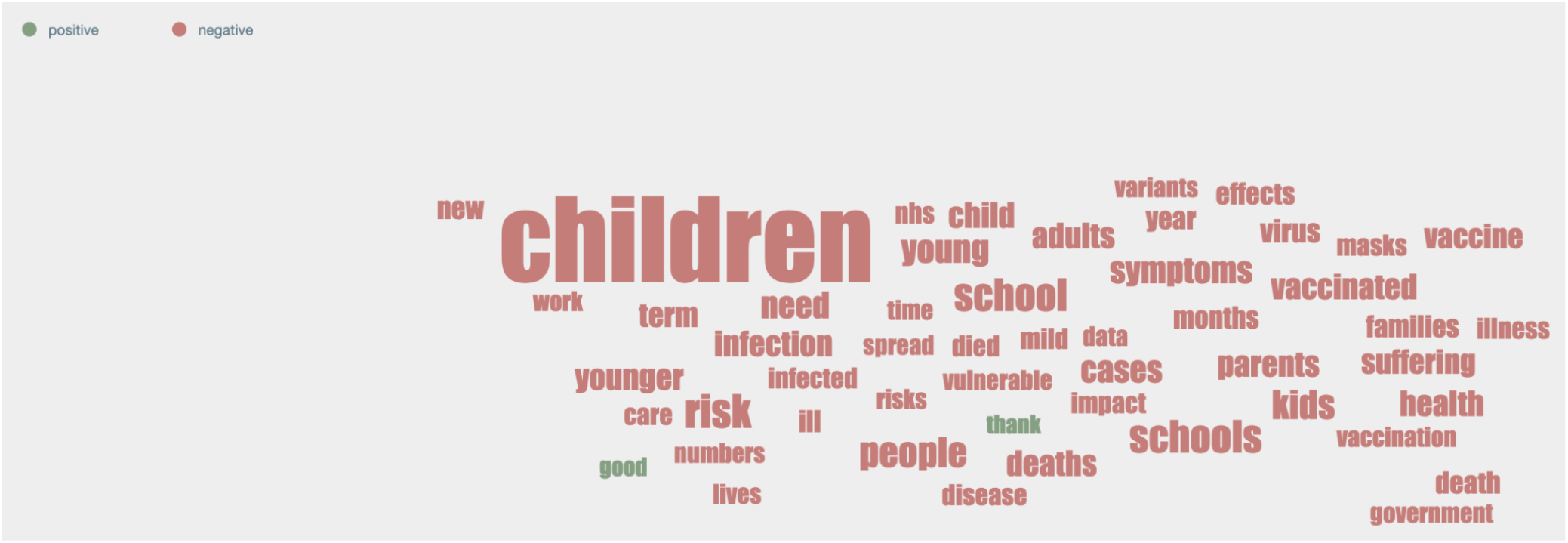
Keywords Word Cloud by sentiment in Long COVID CYP analyses

The primary emotions identified in the word cloud were (Figure 2):

**Figure 2:**
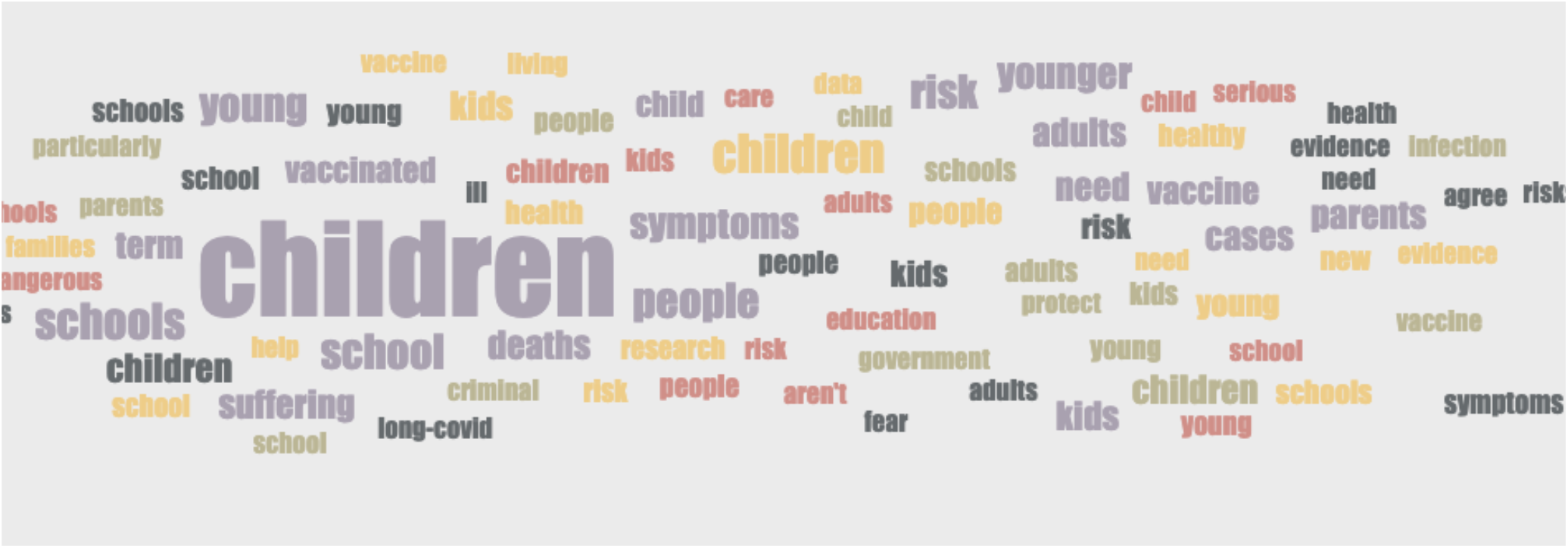
Keywords Word Cloud by emotion in Long COVID CYP analyses

1. Sadness (67.7%; n=3,889), *such as in the following tweet: @Username “After my on-call last night, seeing hospitalisations going up, Gov announcement and people minimising effect of long-Covid in adults and children, it’s very hard to keep spirits up today. We’ll continue to do our best in the NHS”*.
2. Joy (14.2%; n=816): *@Username “What a day! (*…*) I loved sharing my clinical pearls I’ve learned over the past year supporting kids with Long Covid”*.
3. Fear (9%; n=517): *“@Username We urgently need to get the message out that youngsters need to get jabbed, long covid is ruining lives. Currently 1 million in the U*.*K. that’s not 1m with anxiety or hypochondria either. Physiological and measurable changes. Please let us come on to report this”*.

### Segmentation analysis

This analysis revealed the critical clusters of conversation around the main topics of concern within the discourse network around Long COVID in CYP (Figure 3). Comments were distributed in five key conversation segments:

**Figure 3:**
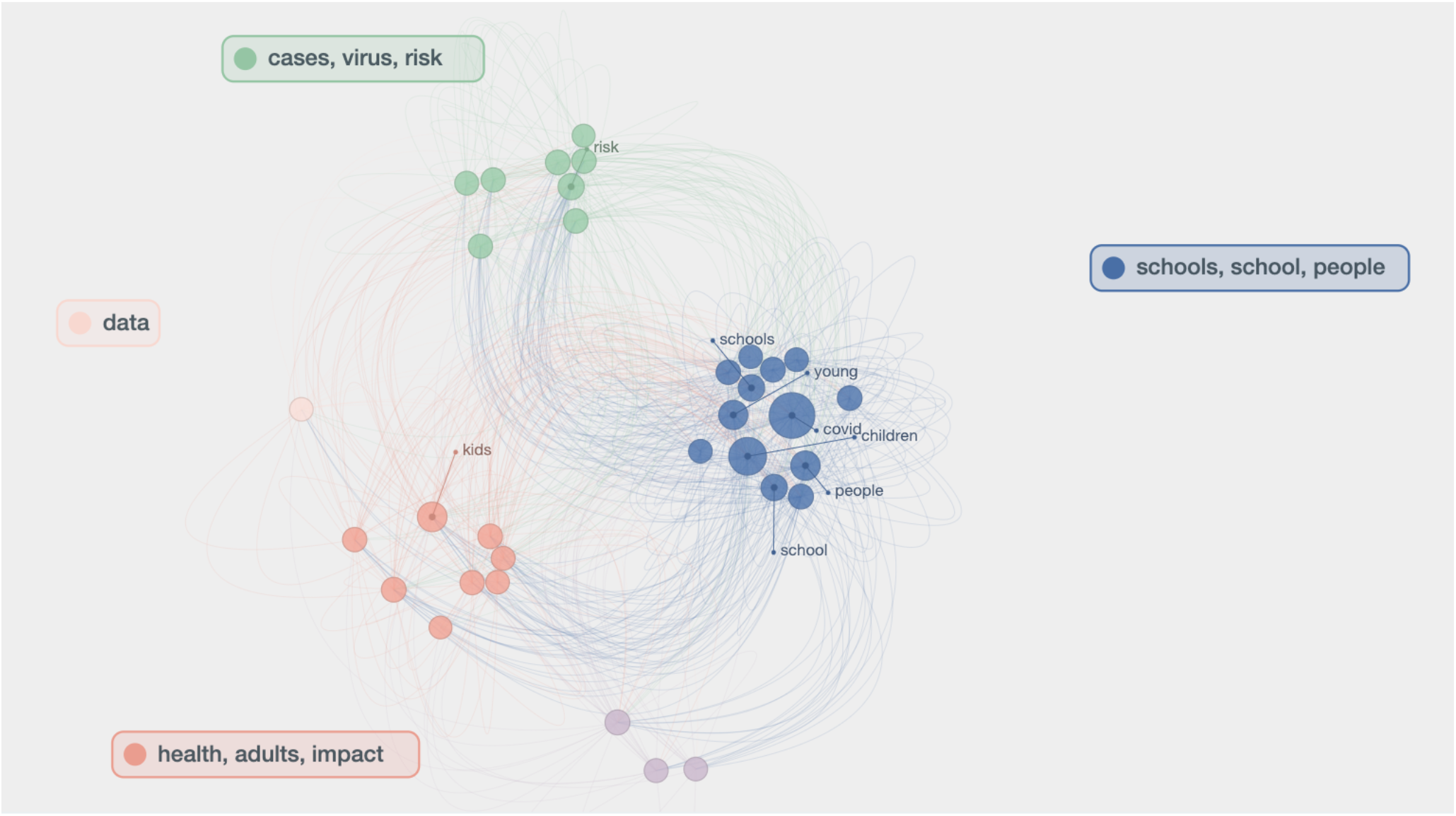
Associations by Keywords, in segments

1. People, schools, school (67%; n=5,239): The measures taken regarding COVID-19 prevention in schools, concern about the risk of exposure, and sharing experiences of infection in schools.
2. Health, adults, impact (15.5%; n=1,212): Reflected concerns and uncertainty about the long-term effect of Long COVID on CYP and adults.
3. Cases, virus, risk (12.6%; n=984): Related to worries about the associated risks and long-term consequences attributable to Long COVID (in both adults and CYP) and the constant mutation of the virus, which will create a permanent risk in the population.
4. Months, suffering, symptoms (4.1%; n=320): Some HCW used Twitter to share how CYP suffer from Long COVID and the extent of these symptoms. Some exemplified some of the typical manifestations, such as fatigue.

### Discourse Analysis by theme

To better understand the topics discussed from the segmentation analysis, we performed a discourse analysis of the key co-occurring themes and topics of concern shared within discussions about Long COVID in CYP. The following themes emerged: concern/uncertainty for the future, school attendance, mask protection from COVID-19, vaccine uptake, infection rates, policy (support/scepticism), understanding and visualising symptoms, child mental health, access to care, community support, and research:

a. Concern for the future/uncertainty (615 tweets, 23.8%): Most comments showed a concern for the future, focusing on shared stats regarding the rate and spread of infection in CYP and how this would affect future health outcomes. This group also expressed concern about political decisions, the presence of illness in loved ones, the eventual overload and response capacity of the health system in the face of an increase in Long COVID cases, and the need for training of HCW to deal with co-morbid, potentially long-term symptoms (Figure 4.1a).
b. Schools (460 tweets, 17.8%): Comments aimed to promote vaccination policies for schoolchildren and flexible measures regarding teachers’ work and attendance, considering cases of people with prolonged symptoms. In addition, several tweets expressed dissatisfaction with school risk mitigation measures, like the use of face masks and air filters, among others (Figure 4.1b).
c. Vaccine (385 tweets, 14.9%): Most of the tweets from this group showed their disapproval of the constant changes in the government’s decisions regarding schools and priority groups for vaccination. Between March and June 2021, the first tweets criticised the lack of priority in the vaccination programme for schoolchildren and other at-risk groups (such as teachers). Once the authorities announced a vaccination programme for 12-15-year-olds (Appendix 1, Table 1), most comments promoted vaccination for this group. A few comments (3%) shared concerns about the vaccine’s efficacy for children, based on experiences of COVID-19 reinfection in adults despite having received the recommended initial doses. However, to a lesser extent (1%) there was a refusal to vaccinate children, citing fear of possible adverse effects. However, the community frequently refuted such comments (Figure 4.1c).
d. Share statistics (334 tweets, 12.9%): Frequently HCW share statistical data, like the number of sick CYP, the number of Long COVID cases, hospital admissions and deaths, among others. Some of this data was used to validate the existence of the Long COVID phenomenon or to express concern about it (Figure 4.1d).
e. Policy (315 tweets, 12.2%): The comments were responsive to the policies emanating from the authorities over time (see Appendix 3). There were five main criticisms: (i) changes in school closure/opening policies, ii) HCW question why the authorities ignore the evidence of Long COVID cases in CYP, leading them to question whether decision-makers have sufficient training to control the pandemic adequately; (iii) the failure to include teachers and school workers in the COVID-19 vaccination program, as well as the younger population; (iv) the lack of mitigation measures in schools, such as improvements in ventilation systems and mandatory use of masks; and (v) herd immunity as a plan in the government’s “hidden agenda”, i.e. “The gov wants ur kids at school so you can work and spend, NOT because it is for your benefit or ‘freedom’…” (Figure 4.1e).
f. “Proof” (279 tweets, 10.8%): Most of the tweets in this group argue for the existence of Long COVID CYP through pictures, statistics, scientific articles, and personal, family, and professional experiences (Figure 4.1f).
g. Signs and symptoms (190 tweets, 7.3%): Among the symptoms described, chronic fatigue and exhaustion are the most frequent, which prevent normal activities. Other symptoms were: i) respiratory: dyspnoea, chronic cough, shortness of breath; ii) gastrointestinal: acute/intense abdominal pain, nausea, bloating, gastroparesis, change in smell/taste; iii) muscular: severe joint pain, “painful foot,” difficulty with physical activity; (v) mental health: anxiety, low mood; vi) topical: rash, skin rashes, redness and pain in the eyes, and finally; v) non-specific symptoms such as chest pain, heart palpitations, constant high temperature, precocious puberty, hormonal changes, and erectile dysfunction (Figure 4.1g).
h. Facemasks (118 tweets, 4.6%): Facemasks were widely promoted, especially in schools, because HCW considered them a practical and straightforward strategy to control the pandemic (Figure 4.2h).
i. Scepticism (102 tweets, 3.9%): Comments showed reticence towards Long COVID in CYP. Some of the arguments focussed on a perceived lack of clarity in the clinical manifestations and stressed a need to better differentiate Long COVID from other related symptomatologies, such as mood disorders (depression, anxiety due to confinement). On the other hand, several agreed on the need for more scientific evidence, arguing that Long COVID in CYP cases are isolated. Other users claimed not to know of such cases, instead of calling the Long COVID in CYP message an exaggeration. Also, several favoured releasing restrictions for CYP, particularly those related to the use of masks, because of possible associated risks, e.g. hypoxia (Figure 4.2i)
j. Mental health (54 tweets, 2.1%): Symptoms attributable to mental health problems in CYP were also a concern. For instance, HCW mentioned sadness, fear of infecting their family, anxiety about sick parents, stress, night terrors, self-harm and suicidal ideation. Also discussed was a perceived lack of specific support for CYP and their families in situations such as hospitalisation, prolonged COVID-19, admission to intensive care, death of a family member, schoolmate, or teacher, all situations that trigger permanent stress in these groups, is also described (Figure 4.2j).
k. Community support/asking for advice (92 tweets, 3.6%): Some HCW used Twitter to ask for guidance on a specific issue or share experiences of having Long COVID or caring for CYP or family members. They also shared informative infographics about Long COVID CYP’s expert opinions (Figure 4.2k).
l. Access to health care/treatment (72 tweets, 2.8%): Some HCW mentioned the lack of specialist (cardiology) support, concerns about prolonged NHS burnout and criticisms about how follow-up was carried out concerning the relative symptomatology of CYP with Long COVID. At the same time, opening new centres for CYP with Long COVID generated different reactions. On the one hand, some HCWs recognised it as a significant development, but on the other hand, as proof of the existence of Long COVID in CYP, which raised concerns for the future (Figure 4.2l).
m. Research (51 tweets, 2.0%): Under this theme, tweets largely promoted current research on Long COVID in CYP or highlighted the need for further research on the subject (Figure 4.2m).
n. Images (58 tweets, 2.2%): Images shared were primarily from scientific studies, including infographics (from organisations such as NHS or @LongCovidKids) and visualisation of CYP’s symptoms, such as rashes, COVID -toe and joint pain. Most infographics shared by organisations (and not individuals), such as the organisation LongCovidKids, were related to statistics, such as the number of CYP with Long COVID or the quantification of the type of symptoms experienced. Shared photographs tended to show the more “visually recognisable” symptoms of Long COVID, such as skin lesions/rashes/inflammation. The less visible symptoms, such as chronic fatigue and neurological issues, were represented with photographs of CYP lying/sleeping under blankets/duvets or on hospital beds (Figure 4.2n).

**Figure 4.1.**
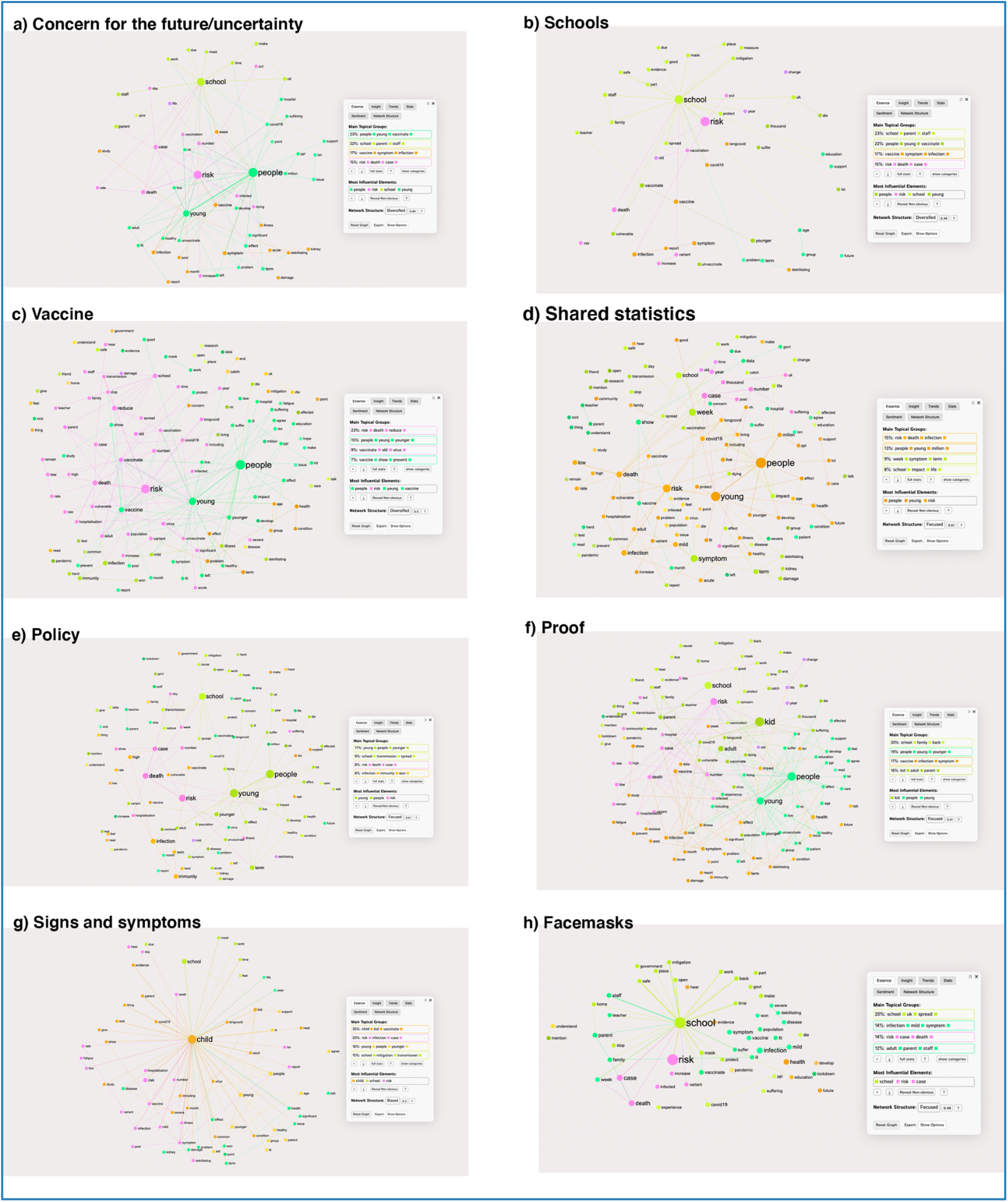
Discourse Analysis by theme

**Figure 4.2.**
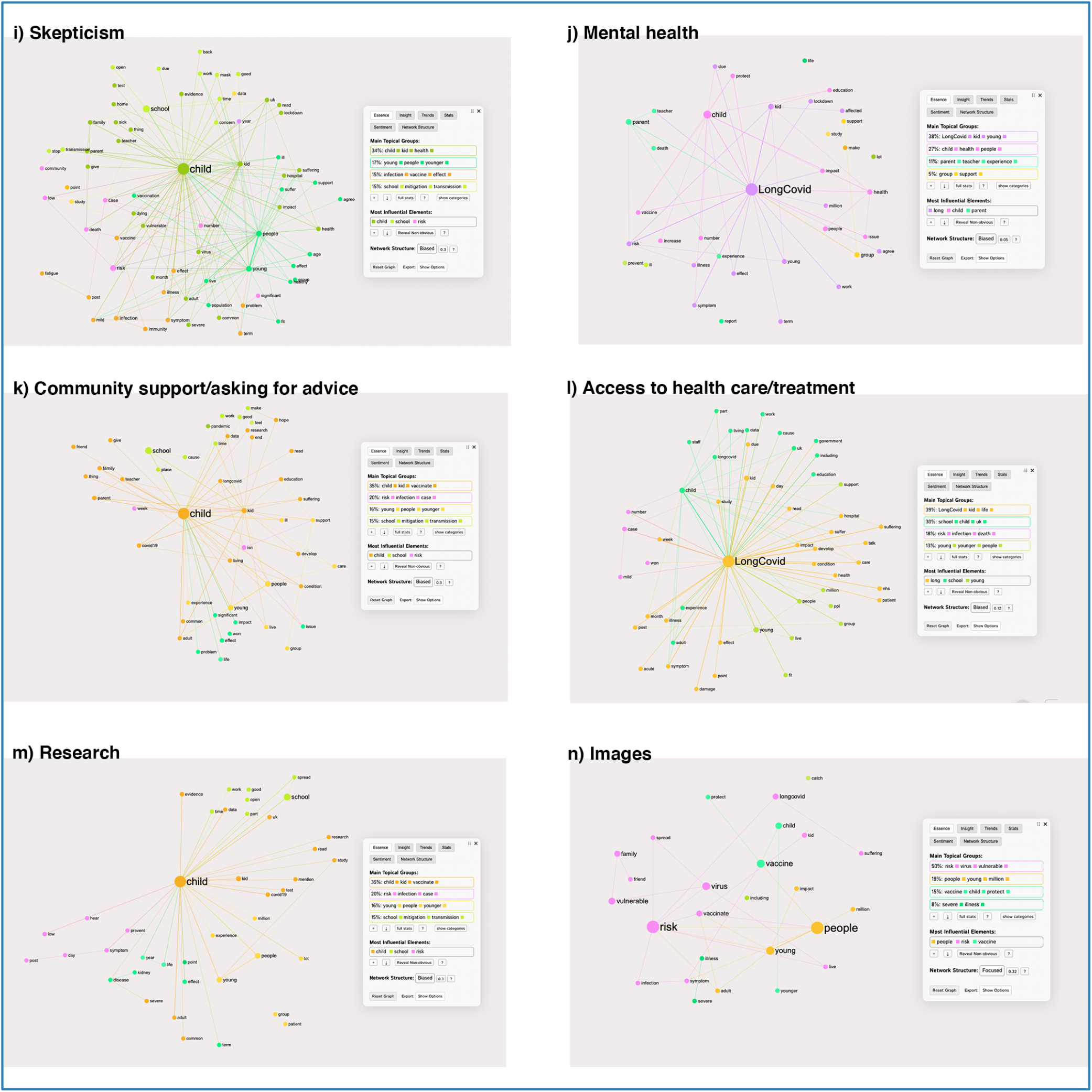
Discourse Analysis by theme

## DISCUSSION

Our primary objective was to explore HCW’s perceptions concerning Long COVID in CYP in the UK between January 2021 and January 2022. Our findings indicated that comments made by HCW on Twitter were responsive to announcements issued by authorities regarding the management of COVID-19 in the UK and associated regulations on the operation of schools. The most frequent feelings and emotions were negative, mainly sadness. In turn, we identified relevant themes for HCW, such as uncertainty or concern about the future, policies and regulations for the prevention, managing and addressing both COVID-19 and Long COVID in CYP, vaccination, and the use of Twitter as a strategy to share scientific literature, management strategies, and clinical and personal experiences.

Concern from HCW about the policies for addressing COVID-19 in the UK in CYP (including vaccination and schools) was a recurring theme in our findings. Concern about the side effects of the COVID-19 vaccine and how the vaccine might interact with pre-existing physiological symptoms of Long COVID in CYP - was also a topic of concern. Likely, the constant change in policy-making in the UK - as public health bodies and governments have tried to understand and adapt to the emergence of Long COVID - has added to the strength of this ongoing debate.^31^ The lack of up-to-date evidence on Long COVID in CYP prompted HCW to rely on using Twitter during the pandemic as a tool to communicate relevant information.^32^ Twitter has a broad audience reach, is used as a communication tool by politicians, health bodies and other key influences, and facilitates real-time updates.^33^ During the pandemic, HCWs, primarily those in frontline roles and local response coordinators, have often been challenged to become credible spokespersons for pandemic information.^34^ Such credibility directly influences public confidence and decision-making, which ultimately determines the success or failure of a public health intervention.^33^

Failures in risk communication could also explain the presence of uncertainty and negative feelings associated with school regulations. When people are upset, distressed or fearful, they often do not trust authority, decrease the perceived validity of the communication received, and information processing becomes difficult.^35^ In this regard, Fotheringham et al.^36^ indicated that during 2020 school leaders in the UK faced pressures and challenges related to translating and enacting school policies, particularly with the perceived lack of agency shared by the government concerning being able to translate centrally issued guidelines. In turn, Tomson et al.^37^ reported that the pandemic has negatively impacted the well-being of leaders in all types of schools and across all demographic groups, affecting their ability to think clearly and solve work-related problems. Given that the protection and care of CYP health during the COVID-19 pandemic ultimately rests with school leaders, the search for support strategies that focus on these groups’ needs becomes an urgent necessity.

### Findings in relation to other studies

This is one of few studies to collect HCW’s perceptions regarding Long COVID in CYP in the UK, using information from Twitter. However, it is not the only study that has addressed Long COVID on this social network. Callard and Peregov^38^ reviewed how, through social platforms such as Twitter, patients made visible the persistence and heterogeneity of COVID-19 symptoms, thus catapulting the inclusion of Long COVID as a relevant phenomenon in clinical and policy discussions. On the other hand, Davis et al.^39^ conducted an online survey on various platforms (including Twitter) to determine the symptoms of Long COVID in people aged 18 and over, seven months after suspicion and confirmation of COVID-19. In turn, Miyake and Martin^40^ reviewed posts mentioning long-term symptoms of COVID-19 on social platforms such as Twitter, Facebook, blogs, and forums. The authors identified time/duration, symptoms/tests; emotional impact; lack of support and resources as critical issues. It was also reported that due to conflicting guidelines and rule breaches by politicians themselves - throughout the pandemic, that the general population in the UK had decreased levels of trust in the UK government as a source of information about the coronavirus and a decreased trust in politicians and news organisations. In the same context, Enria et al^35^ showed that people with higher levels of education were less likely to have a favourable opinion of government decisions.

### Implications for policy and practice

Several policy recommendations and implications are targeted at various stakeholders to consider when implementing future policy guidelines to address Long COVID healthcare delivery. First, policymakers should consider investing appropriate resources to collect data regarding Long COVID in CYP, specifically around the impact of COVID-19 on the mental health of CYP. The above implies working closely with researchers to streamline data collection and reporting on Long COVID. Second, policymakers should consider providing a basic level of psychosocial support with access to quality mental health and psychosocial support services for HCW, school staff, parents, and CYP suffering from Long COVID. The above implies strengthening health systems, community-based programming and mobilisation. Policies must include documenting the impact of MHPSS interventions and innovative approaches to be more widely disseminated and scaled up across different contexts and target population groups. Third, to address the criticism around frequent changes in school closure and opening policies, decision-makers should develop clear and easy-to-understand school mitigation plans informed by the best available evidence. The plans should incorporate teachers, school workers, and parents to ensure that all voices are included in the policy plan. Fourth, policymakers should adopt a shared decision-making approach incorporating HCW in the decision-making process for managing COVID-19. Lastly, government decision-makers should set Long COVID pandemic recovery policies that are informed from a health equity perspective and how this affects CYP living with Long COVID - factoring in childhood, family income, housing, domestic violence, access to healthcare, and racism.

### Strengths and limitations

A key strength of this study is that our online analysis of Long COVID contributes towards an emerging understanding of reported experiential, emotional and practical dimensions of Long COVID in CYP specifically, as well as questions of vaccine hesitancy in CYP with Long COVID. This is one of few studies to collect HCW’s perceptions regarding Long COVID in CYP in the UK, using information from Twitter. We identify key areas that need considering attention and focus, such as the provision of psychosocial support with access to quality mental health resources to alleviate the impact of Long COVID in CYP; and the development of clear Long COVID pandemic recovery guidelines that are informed by a health equity perspective and how this affects CYP living with Long COVID. One of the limitations this study acknowledges is the definition of Long COVID in CYP. When data were collected, the lack of consensus on the definition of Long COVID in CYP forced us to formulate a definition of Long COVID in CYP based on the available literature. Furthermore, this study is limited to the perceptions of people who used descriptors in their online biography attributable to HCW, so our results are not representative of all HCWs in the UK nor of HCWs in other countries. Nor do they represent the perception of Long COVID in CYP in the general UK or the global population. In turn, this research collected data from Twitter only, so further inquiry into HCW perceptions of Long COVID in CYP required expansion to other data sources or social networks and inclusion of languages other than English.

## CONCLUSION

More than a year after the start of the COVID-19 pandemic, the perceptions described on Twitter by HCW concerning the presence of Long COVID in CYP appear to be a relevant and timely issue, very responsive to the declarations and guidelines issued by health authorities over time. The most prominent group within the discourse studied was the activist/lobbying organisation: @LongCovidKids, which shared the most tweets and images over the period studied. We recommend that future research focus on how online health activism is organised and carried out for CYP with Long COVID. We believe that such a strategy would allow for a better understanding of the scope and impact of this phenomenon and how it can influence decision-making. Also, we suggest different mitigation strategies, support and training of HCW and school staff - regarding manifestations and treatment of Long COVID in CYP across all demographic areas.

## Data Availability

Data included in this study is not available outside the Rapid Research Evaluation and Appraisal Lab (RREAL), UCL due to ethical guidelines restrictions

## Acknowledges

We are grateful to the Rapid Research Evaluation and Assessment Laboratory (RREAL), Department of Targeted Intervention, University College London, London, UK, whose support has been essential for the development of this project.

## AUTHOR STATEMENT

- **Martin, Sam:** Made substantial contributions to the conception and design, analysis, and interpretation of data for the work. Also, she drafted the work and revised it critically for important intellectual content. In addition, she gave the final approval of the version to be published; AND she agreed to be accountable for all aspects of the work in ensuring that questions related to the accuracy or integrity of any part of the work are appropriately investigated and resolved.
- **Chepo, Macarena:** Made substantial contributions to the conception and design, analysis, and interpretation of data for the work. Also, she drafted the work and revised it critically for important intellectual content. In addition, she gave the final approval of the version to be published; AND she agreed to be accountable for all aspects of the work in ensuring that questions related to the accuracy or integrity of any part of the work are appropriately investigated and resolved.
- **Deom, Noemie:** Made contributions to the analysis AND revised it critically for important intellectual content, AND she gave final approval of the version to be published, AND she agreed to be accountable for all aspects of the work in ensuring that questions related to the accuracy or integrity of any part of the work are appropriately investigated and resolved.
- **Khalid, Ahmad Firas:** Made contributions to the analysis AND revised it critically for important intellectual content, AND he gave final approval of the version to be published, AND he agreed to be accountable for all aspects of the work in ensuring that questions related to the accuracy or integrity of any part of the work are appropriately investigated and resolved.
- **Vindrola-Padros, Cecilia**: Made contributions to the analysis AND revised it critically for important intellectual content, AND she gave final approval of the version to be published, AND she agreed to be accountable for all aspects of the work in ensuring that questions related to the accuracy or integrity of any part of the work are appropriately investigated and resolved.

