## Appendix for "“#LongCOVID affects children too”: A Twitter analysis of healthcare workers’ sentiment and discourse about Long COVID in children and young people in the UK"

**Appendix 1**

Table 1. Timeline of national governmental policies/guidelines regarding CYP (Long Covid and schools). Sources: Official information from pages of the NHS,[40] [2] Gov.UK,[41]and press.[42,43]

| Date of (first) publication | Name / Title | Source | Target Audience |
|---|---|---|---|
| 27/04/2020 | COVID-19: paediatric surveillance | PHE | Healthcare Providers |
| 01/2021 | Schools closures are announced | | |
| 03/2021 | Reopening of school | | |
| 03/2021 | Face coverings are required in schools and colleges | | |
| 04/2021 | Face coverings no longer required in schools and colleges | | |
| 05/06/2021 | Long Covid plan 2021/2022: 10 key next steps to support those suffering from Long Covid | NHS | Healthcare Providers |
| 09/2021 | Covid-19 vaccination programme extended to 12-15 years old | | |
| 02/09/2021 | First findings from world's largest study on Long Covid in children and young people (cites the: Non-hospitalised Children & young people (CYP) with Long Covid (The CLoCk Study)) | NIHR | Healthcare providers |
| 11/2021 | Face coverings are required in schools and colleges | | |
| 12/2021 | School closures are announced | | |
| 01/2022 | Reopening of schools | | |
| 01/2022 | Face coverings no longer required in schools and colleges | | |
| 02/2022 | End of mask-wearing policy | | |
| 02/2022 | Covid-19 vaccination programme extended to 5-11 years old | | |

Table 2: Filters used for the search strategy on Twitter

| Variable | Descriptor |
|---|---|
| Keywords | kid* OR child* OR school* OR young* OR parent OR parents OR toddler* OR nursery OR kindergarten OR boy OR girl OR teen OR teenager OR infant OR infants OR minor OR underage OR "primary school" OR "secondary school" OR classroom OR playground |
| Bio | nurs* OR physio* OR physici* OR "midwife" OR "obstetrician" OR geriatric* OR "HCW" OR "health care worker" OR "healthcareworker" OR "doctor" OR "GP" OR "general practitioner" OR "health care professional" OR "health professional" OR pharmac* OR radiograph* OR therap* OR "neurologist" OR psy* OR clinic* OR "ambulance" OR "NHS" OR "osteopath" OR "orthoptist" OR "immunologist" OR "oncologist" OR "endocrinologist" OR "gastroenterologist" OR gastro* OR "medical" OR "medic" OR "audiologist" OR "sonographer" OR cardio* OR paediatric* OR pediatric* OR "dietitian" OR "paramedic" OR "pathologist" OR anaesth* OR anesth* OR "orthoptist" OR surg* OR "dermatologist" OR "dentist" OR "geneticist" OR "rheumatologist" OR "urologist" OR "haematologist" OR "neonatologist" OR intern* OR "podiatry" OR "neurosurgeon" NOT "lecturer" NOT "academic" NOT "professor" |

**Appendix 2**

**Theme Codebook : examples of tweets that fit into main themes tagged for mention of CYP with Long COVID**

| Tag | When to apply | Example | Classification of the example according to the type of feeling |
|---|---|---|---|
| LCK – call for advice | Request advice on how to deal with LCK (symptoms, consultations, access to services, etc.) | Looking for private doctor to diagnose / treat suspected encephalitis in a boy that had covid last year. London-based, but telemedicine would be fine. Accelerating deterioration of focus over past two months. Suggestions please? #LongCovidKids #LongCovid | Negative ☹️ |
| LCK – call for more support | Request more support from formal agencies (NHS). | #LongCovidKids - received a text this am. My son is 17, final year at school but he had a positive test on August 14th and now has long COVID. He has barely attended this term as he feels so unwell and exhausted. Little nhs support. Sorry to ask but do you have any advice… 1/ | Negative ☹️ |
| LCK- community support | Referred to expert support, support groups of parents, friends or relatives, among others. | We welcomed 104 new families to our support group yesterday. Some with more than one child living with #longcovid<br><br>Over the coming week they will share their experiences & our volunteers will try to guide them while others chose to minimise.<br><br>#LongCovidKids | Positive ☺ |

| | | | |
|---|---|---|---|
| LCK-facemasks | Discuss use of face mask as a pandemic control | A well fitting mask, good quality mask 😷 is the one defence young children have.<br><br>Avoid mixing indoors with others, especially where masks are not worn.<br><br>We don't know which of these children will develop #LongCovid<br><br>14% could.<br>#MaskUp #BackToSchool #LongCovidKids | Positive<br>☺ |
| LCK-family concern | The comment alludes to their experience with LCK as a parent, familiar, friends, among others | My nephew has covid for the 2nd time from school. He's trying to study for his GCSEs, his mum is clinically vulnerable, his dad has long covid from last time you failed to put mitigations in place. I will never forgive your government for your blatant disregard for us all | Negative<br>P |
| LCK – expert opinion | Rrefers to an expert opinion, whether in a tweet, talk, lecture or otherwise | What a day at the @*Username* Long Covid conference lecturing alongside Dr Tina Peers & Robyn Puglia - they called us "The Three Musketeers" - I loved sharing my clinical pearls I've learned over the past year supporting kids with Long Covid<br><br>#LongCovidKids #longcovid | Positive<br>☺ |
| LCK-health access | Refers to access to and use of health services (diagnosis, treatment, follow-up, acceptability, etc.) | @@*Username* @@*Usernames*<br>There is no magic NHS staff tree. "Creating capacity"is a non starter.<br>We are under doctored, under nursed, under midwifed.<br>There are vacancies throughout the system, and that was before accounting for staff isolating, | Negative<br>☹ |

| | | | |
|---|---|---|---|
| | | caring for sick kids, Long Covid and burnout. | |
| LCK – home organization | Discuss changes in household organisation due to covid | @@*Username*Thanks for that. Unfortunately I'm suffering from long COVID and trying to run a business while bringing up 3 kids so forgive me if I need the odd mindless distraction. | Negative ☹️ |
| LCK- images | A picture of a symptom, relevant event, health centre, among others, is shown. | Photograph of child and visual rash (image redacted for ethics) | Negative ☹️ |
| LCK – is real | Activism about the existence of Long Covid in kids, and rejection of denialism or minimisation of it | *@Username@Username@Username@Username* Children are loosing their lives to Covid and being left with long Covid too you know | Negative ☹️ |
| LCK- mental health | Alludes to impact on mental health (distress, sadness, stress, depression, uncertainty). | Group Post<br><br>“My teen D just broke down 😢 shes had thoughts about self harm. She can’t take #LongCovid anymore, she feels empty. She has been keeping it from me as she didn’t want to bring me down as I’m still ill too. We are so broken and I can’t help her”<br><br>#LongCovidKids | Negative ☹️ |
| LCK- need for HW training | Need for education or further training of health workers for different long | *@Username@Username@Username* We aren't taught about it at all in med school or GP training. Many colleagues have never heard of it. I only learnt about GET being bad from a pt. | Negative ☹️ |

| | | | |
|---|---|---|---|
| | covid treatment/therapies | I only know about any of this now through my own illness w #LongCovid<br>We need to lobby the med schools- not hate the practitioners | |
| LCK – policy | Refers to formally issued directives or guidelines. | In the last 3 weeks, 10,000 infections a day amongst 5-14 yr olds.<br>Literally countless Long Covid cases (likely many tens of thousands).<br>In the autumn term so far nearly 1,000 6-17 yr olds hospitalised.<br>A national crisis!<br>So what is the Government doing to protect children? | Negative 🙁 |
| LCK - research | Call to generate or disseminate more research in long covid | Paediatric follow up open protocol & data collection forms available from *@Username* for independent & collaborative global studies. Sites globally welcome to Join *@Username* follow up working group (adult & children) #LongCovid #LongCovidKids | Neutral |
| LCK- School | Tells of LCK's experience in schools (contagion, non-attendance) | By everybody, you mean adults. Teens are not boosted. Small children are not vaccinated. Schools have little protection. Learning to live with covid means accepting our children get it repeatedly, many will have longcovid, & be disabled & some will die. | Negative 🙁 |
| LCK- scientific article data | Publications or data from scientific articles or other formal studies are shared. | The government's own figures (from ONS) say 8% of infected kids experience #LongCovid. That could be several hundred thousand children.<br><br>You don't want that to happen to your children, right? We don't know what the long-term effects are. For all we know, they could be lifelong. | Negative 🙁 |

| | | | |
|---|---|---|---|
| LCK - symptons | Describes presence of symptoms attributable to LCK | Group Post<br>“I’m really battling atm with school. 8 yo hasn’t been concretely diagnosed with #LongCovidKids – they cant find a cause for continual tummy pain since May, nausea, fatigue, low mood, now anxiety cause it’s gone on so long”<br>This is living with #COVID19 #LongCovid | Negative<br>☹ |
| LCK – treatment | Describes the use of any strategies to treat symptoms of long covid in children | CoP has year of experience in managing & delivering care for people of all ages, incl children, with #LongCovid in community & primary care. Learning from emerging evidence plus EbEs. Members would love to share their learning & experience of emerging models | Positive<br>☺ |
| LCK – uncertainty | Refers to uncertainty regarding symptoms, diagnosis, prognosis. | Do we know how common long covid is in children? | Neutral |
| LCK-vaccine | Call or concern about vaccination in children and adolescents | Vaccs reduce transmission.Even Delta (but not as well as for other strains). Kids need protecting - over 100 died of Covid in UK,100s in hospital with COVID and 1000s living with long covid. The vaccine is tested. It is less risky than getting immunity from infection.Masks work | Negative<br>☹ |
| LCK - videos | A video of a symptom, relevant event, expert opinion, among others, is shown. | - | - |

**Appendix 3**

**Sentiment analysis framework: Attitudes towards Long COVID in CYP**

**Positive (P)**

- Post communicating overall trust and satisfaction with public health guidelines and support for treatment of Long COVID in CYP.
- Posts are affirming of official policies regarding the treatment of Long COVID in CYP.

**Negative (N)**

- Post contains negative attitudes/arguments against public health guidelines and support for treatment of Long COVID in CYP.

- Post discourages the following of recommended guidelines/support related to treatment of Long COVID in CYP - (for personal, political or other reasons).

Post shares bad experiences of treatment of Long COVID in CYP.
